# Randomized Double Blind Inpatient Study of a Gluten-Free Diet in Persons with Schizophrenia

**DOI:** 10.1101/2025.02.24.25322813

**Authors:** Deanna L. Kelly, Christopher M. Lee, Daniel J.O. Roche, Monica V. Talor, Sarah Clark, William W. Eaton

## Abstract

**Background:** Schizophrenia and related disorders (SRD) are characterized by positive and negative symptoms, such as anhedonia and avolition. There are no current FDA approved treatments for negative symptoms, which is a critical gap in our treatment of people with SRDs, since they are a major determinant of functional impairment. An emerging literature suggests that SRDs have a relationship with immune function and inflammation. Recently an SRD subgroup with high inflammation and elevated levels of anti-gliadin antibodies (AGA) of immunoglobulin G type (IgG) ha**s** been characterized. Negative symptom improvement has been previously observed with gluten removal in this subgroup in two small clinical trials.

**Methods:** We conducted a 5-week confirmatory double-blind placebo-controlled trial of a gluten free diet (GFD) versus gluten-containing diet (GCD) for negative symptoms in people with SRD who have elevated AGA IgG (NCT03183609). Participants were between the ages of 18-64 years, had baseline negative symptoms and a diagnosis of schizophrenia or schizoaffective disorder. Those included were screened for an AGA IgG >20 U, no serologic evidence of celiac disease, and stable antipsychotic treatment and dose. All participants were inpatients, received a GFD and were randomized to 30 grams of gluten or rice flour daily delivered in protein shakes. The Clinical Assessment Interview for Negative Symptoms (CAINS) Motivation and Pleasure (MAP) scale was the primary outcome measure. We also examined the CAINS Expressivity (EXP) scale, the Scale for the Assessment of Negative Symptoms (SANS), the Brief Psychiatric Rating Scale (BPRS), the MATRICS Consensus Cognitive Battery (MCCB) and conducted regular side effect screening and laboratory measures for safety.

**Findings:** Between 2018 and 2024, we included 39 participants (N=21 GFD and N=18 GCD). There was a significant improvement over time in the CAINS MAP (treatment X time *df*=30.1, *F*=2.78, *p*=0.045) in the GFD compared to GCD, but no significant change in the CAINS EXP, the SANS, BPRS or MCCB. The diet was well tolerated; the most frequently occurring side effects were constipation (38.1% GFD, 33.3% GCD), sedation (33.3% GFD, 50% GCD), dry mouth (33.3% GFD, 33.3% GCD), headache (33.3% GFD, 27.8% GCD), and insomnia (33.3% GFD, 27.8% GCD).

**Interpretation:** This is the first large scale double-blind randomized clinical trial in SRD with AGA IgG+. This replication of smaller studies suggests that negative symptoms, particularly anhedonia and avolition may be improved. However, we did not replicate our previous finding of cognitive improvement and COVID-19 likely impacted the extent of improvement in negative symptoms due to quarantines and lockdowns. More work is needed to determine the mechanism of action of gluten removal in this subgroup with hopes of developing new treatment targets for motivational deficits of this illness.

## Introduction

Schizophrenia and related disorders (SRD) are characterized by positive and negative symptoms, such as anhedonia and avolition. There are currently no FDA-approved medications to treat negative symptoms, even though they are a major determinant of social and occupational function (1) and the most significant predictor of poor long-term one-year outcomes (2). Multiple studies support a relationship between SRD, immune function and inflammation (3-18). Importantly, rates of SRD are higher in the setting of autoimmune disorders(19), as well as inflammatory states of the gastrointestinal tract based on *in vivo* (20) and ex vivo studies(21).

The role of wheat consumption as a potential etiological agent has garnered attention in SRD research. In several landmark studies, rates of SRD tracked closely with rates of wheat consumption during World War II(22, 23); meanwhile, SRD was almost nonexistent in countries with no grain consumption(24-26). In early clinical trials (1950-80’s), the removal of wheat from the diets of people with SRD resulted in mixed levels of symptom improvement(27-34). The lack of consistency of study findings may result from the unavailability of biomarkers to define subgroups of SRD(35-37). The field of psychiatry tended to pursue a monocausal etiology for SRD and other psychiatric illnesses(38) rather than conceptualizing them as syndromes comprised of multiple etiologies(39).

Recent SRD research has identified a subgroup of people with schizophrenia who mount an immune response to gliadin, a protein found in certain dietary grains (wheat, barley, rye, triticale). Defined as having high levels of anti-gliadin antibodies (AGA) of immunoglobulin G type (IgG), this AGA IgG+ subgroup is comprised of 33-38% of persons with schizophrenia, a rate that is three-fold higher than found in healthy controls (40-46). The AGA IgG+ subgroup is characterized by different patterns of clinical symptoms, presenting with greater negative symptoms (43). Furthermore, their negative symptoms may improve after removal of gluten from the diet, based on an open-label (47) and small randomized double-blind study (48). One theoretical mechanism involves the kynurenine pathway, which metabolizes tryptophan. Levels of kynurenine and kynurenic acid (KYNA) not only were elevated in the AGA IgG+ subgroup, but also were lowered after removal of gluten from the diet (49). Other studies have also found elevated kynurenic acid in schizophrenia (50).

Here, we present the results of a five-week double-blind placebo-controlled trial. The aim of this trial was to confirm the efficacy of a gluten free diet (GFD) versus a gluten-containing diet (GCD) for improving negative symptoms in persons with SRD and high AGA IgG.

## Methods

### Study design

The study was a 5-week, double-blind, randomized clinical trial conducted at the Maryland Psychiatric Research Center (MPRC), University of Maryland School of Medicine, Maryland, USA. Screening was performed at the MPRC, its affiliated sites, and Johns Hopkins University School of Medicine, Maryland, USA. The University of Maryland Baltimore institutional review board approved all study activities (HP-00075175). The study was reviewed annually by a data safety and monitoring board and was registered in ClinicalTrials.gov (NCT03183609).

### Participants

All participants provided written informed consent after passing the Evaluation to Sign Consent (with score > 10)(51). Gender was self-reported from the following options: female, male. Over the course of 1-2 screening visits, participants’ psychiatric diagnoses were confirmed by the Structured Clinical Interview for Diagnosis of DSM-IV/DSM 5 (SCID)(52), chosen for its good reliability and validity(53, 54). Medical eligibility was established based on a medical history and physical examination by a medically accountable physician, in addition to a standard blood chemistry panel, complete blood count, urinalysis and electrocardiography. Serum levels of AGA IgG, AGA IgA, tTG were collected using INOVA first generation screening kit. Existing gastrointestinal disorders and dermatological disorders were evaluated, since gluten sensitivity may be related to both.(55)

Women and men (ages 18-64 years) were eligible for participation based on the following inclusion criteria: (1) diagnosis of schizophrenia or schizoaffective disorder defined by DSM-IV-TR criteria and confirmed by the SCID; (2) AGA IgG level > 20 U (AGA IgG+); (3) treatment with the same antipsychotic for at least 4 weeks prior to the study; (4) a Scale for the Assessment of Negative Symptoms (SANS)(56) total score ≥ 20, OR, a SANS avolition or asociality score of ≥ 3.

Participants were excluded based on the following criteria: testing positive for tTG (so that their presumed celiac disease could be appropriately treated); adherence to a gluten-free diet prior to the study; active pregnancy or lactation; history of organic brain disorder or intellectual disability; history of a medical condition whose pathology or treatment could alter the presentation or treatment of schizophrenia or significantly increase the risk associated with the proposed treatment protocol; gluten ataxia determined by physician with aid of Brief Ataxia Rating Scale(57); and DSM-IV/DSM 5 criteria for alcohol or substance abuse (other than nicotine) within the last month.

### Randomization and Masking

Participants, researchers and clinical team members were blinded to intervention assignment. Treatments were assigned at random, using computer-generated permuted block randomization sequences with randomly varied block sizes to limit imbalance in the number of patients assigned to each group, while making it difficult for staff to predict what treatment patients were receiving.

### Procedures

All enrolled participants were inpatients on the research unit (Treatment Research Program at Maryland Psychiatric Research Center/Spring Grove Hospital Center). Participants received 10 g of either rice flour (treatment; Bob’s Red Mill®) or gluten flour (control; Bob’s Red Mill®) mixed in a shake each afternoon that contained water, ice, protein powder and optional syrup of the participants’ choice to make a protein shake (Sunwarrior Plant-Based Protein). The research staff ensured that the entire shake was ingested. Throughout the study, participants in both groups received a gluten-free meal plan, consisting of 21 days of meals in a rotating schedule prepared according to the strict gluten-free operating procedures of the hospital kitchen. The target was to keep the ingestion of gluten below 15 mg per day for those in the active treatment group(58). During the day, a 1:1 staff-to-participant ratio was used to ensure protocol adherence. All packaged food consumed was certified gluten-free.

The study intervention was added to participants’ ongoing antipsychotic regimen. Study physicians were instructed to avoid changing doses of other somatic and psychotropic medications during the study. Anticholinergic medications for extrapyramidal side effects (e.g., benztropine and diphenhydramine), propranolol for akathisia and benzodiazepines for anxiety or agitation (e.g., lorazepam) could be prescribed as needed.

### Outcomes

Raters were required to show agreement with an intraclass correlation coefficient (ICC) >0.8 as compared to gold standard ratings of training tapes and regularly showed ICCs among raters>0.8 in monthly training exercises throughout the course of the study.

## Primary outcome

The primary outcome of this trial was primary negative symptoms at week 5 using the Clinical Assessment Interview for Negative Symptoms (CAINS) Motivation and Pleasure (MAP) and Expressivity (EXP). We also used the SANS(56, 59) total score for the evaluation of negative symptoms which was calculated by subtracting the individual items of inappropriate affect, poverty of content of speech, and attention (SANS-AA; an adaptation previously published).(60, 61).

### Secondary clinical symptom outcomes

The following secondary outcomes were each assessed at baseline, and weekly thereafter. The Brief Psychiatric Rating Scale (BPRS)(62) total score was used to measure global psychopathology and the BPRS conceptual disorganization, suspiciousness, hallucinatory behavior and unusual thought content(63) were used to measure positive symptoms. The BPRS hostility(?) factor was used to measure symptoms of hostility. The Calgary Depression Scale (CDS)(64) total score was used to measure depression; the CDS was chosen because it was designed explicitly for schizophrenia(65), outperforms other depression scales in schizophrenia(66), is related to outcome and quality of life, and has reference values for the general population(67, 68). Global Severity of Illness was measured using Clinical Global Impression Scale (CGI).(69) Executive function was assessed using the Brief Assessment of Cognition in Schizophrenia Tower of London Test (1, 70).

The following were assessed at baseline and at study completion. Cognition was assessed using the Measurement and Treatment Research to Improve Cognition in Schizophrenia (MATRICS) Consensus Cognitive Battery (MCCB)(2), both as individual domains and as a composite score of all seven domains standardized to the general population.(2, 71) General Health and Well-being was assessed using the Short Form-36(72).

### Laboratory monitoring and side effects

To monitor general medical health, each participant completed the following at baseline and upon study completion: a medical history and physical examination; routine bloodwork; the Neurological Evaluation Scale (NES)(73); and the Brief Pain Inventory (BPI)(74). Each week throughout the study, participants completed vital sign assessments; the 15-item Gastrointestinal Symptom Rating Scale (GSRS)(75); the Simpson-Angus Extrapyramidal Symptom Rating Scale (SAS)(76); the Barnes Akathisia Scale (BAS)(77, 78); and the 23-item Side Effect Checklist (SEC). Other spontaneous adverse events were also recorded. Body mass index (kg/m2), complete blood count, and complete metabolic panel 14 item were also assessed.

### Secondary laboratory outcomes

Blood samples were collected at baseline and study endpoint for all laboratory tests. For analysis of AGA IgG, serum separator tubes (SST; tiger top cap) were drawn and centrifuged immediately. Approximately 6-8 mL sera (in 4-5 aliquots of 1-2 mL each) were aliquoted into 3.5 mL twist top freezer tubes, stored in a -80°C freezer, then delivered to the Cihakova lab at Johns Hopkins University on dry ice for cytokine assessment.

Additionally, AGA IgG was measured and analyzed using semi quantitative ELISA assays from Inova Diagnostics (catalog # 708650). Cytokines were measured using EMD Millipore’s MAP Human Cytokine Magnetic Bead panel (Luminex bead-based immunoassays (Millipore, Billerica NY). The readout was completed using a Bioplex 200 platform (Biorad, Hercules CA) to determine the concentration of multiple target proteins in the specimens.

#### Statistical analysis

All analyses were performed on the modified intention-to-treat principle, according to randomized treatment assignment. The group-level difference between treatment groups for outcome measure scores obtained at study endpoint were evaluated using t-sample t-test; and Cohen *d* was calculated for any comparison that reached significance. The effect of treatment on rate of change for each outcome measure was calculated using least squares mean, in which the group mean was calculated from the following analysis of covariance (ANCOVA) model: change from baseline = baseline + treatment + week + treatment x week. This *a priori* model served to control for covariates, including baseline differences in the presentation of the data.

For each biological outcome, the effect of GFD versus gluten containing diet on each outcome measure was estimated by fitting the ANCOVA model: endpoint measure = baseline value + treatment.

#### Choice of primary measure

CAINS MAP was selected as the primary outcome measure for this study examining negative symptomatology in SRD. The CAINS offers advantages over other commonly used measures of psychotic symptoms, such as the SANS, Positive and Negative Symptom Scale (PANSS), or the Brief Negative Symptoms Scales (BNSS). Advantages include: (1) the incorporation of both observed and subjective report, (2) avoidance of symptoms relevant to other psychopathological dimensions of functioning, and (3) providing separate assessment of consummatory vs anticipatory anhedonia (85). During a pandemic the inclusion of observed measures and anticipatory features of anhedonia were important to optimally study negative symptoms. The CAINS is widely administered with over 80 citations in PubMed, it has been translated into several languages, and it was empirically developed and evaluated to measure negative symptoms in SRD. Psychometric properties include good internal consistency, test-retest stability and interrater agreement. It has been validated against other negative symptoms scales related to pleasure and motivation and discriminant validity shows it is independent from cognitive measures, depression and medication side effects. In fact, the CAINS scales were also designed to map onto functional outcomes. A full manual and scoring are available.

## Results

From July 10, 2017, to March 30, 2024, a total of 41 participants were randomized, and 39 participants met *a priori* criteria for study inclusion with at least one full week of data. There were no significant differences in baseline demographics or characteristics 1. Seven participants withdrew from study participation due to the COVID-19 outbreak (N=1, controls), stomach discomfort (N=1, controls), not enough food (N=1, controls), COVID infection (N=2, GFD), or were discharged from the inpatient unit (N=1, controls; N=1 GFD).

### Psychiatric Symptoms

Based on ANCOVA models, there was a significant main effect of treatment on change in CAINS motivational deficits (MAP) (*df*=30.1, *F*=2.78, *p*=0.045), such that participants treated with GFD had greater rate of symptom improvement over the 5-week trial. There was a trend-level effect of treatment on change in SANS anhedonia/asociality (*df*=29.5, *F*=1.84, *p*=0.15), with the GFD group showing greater symptom improvement. There were no significant effects of treatment on change in symptom severity for any of the other symptom outcome measures, including CAINS expressive deficit (*p*=0.87), SANS total (*p*=.57), SANS blunting (*p*=0.79), BPRS total (*p*=0.18), CDS (*p*=0.62), CGI-severity (*p*=0.91), or CGI-global (*p*=0.42). There was a trend for BPRS positive symptom improvement (p=0.08) in the GFD relative to GCD.

There were no significant effects of treatment on change in cognitive symptoms, including the MCCB total or domain scores (all *p values >* 0.43), or the Tower of London (*p*=0.17). Similarly, there were no significant effects of treatment on change in health or gastrointestinal symptoms, including the Short Form-36 total or domains (*p*=0.29), NES total or domains (*p*=0.35), the GSRS total or domains (*p*=0.34), BPI-severity (*p*=0.79), or the BPI-impact (*p*=0.52).

#### Laboratory Measures

There was a significant effect of treatment on change in IL-2 levels (*df*=27.9, *F*=3.5, *p*=0.019), such that participants treated with GFD experienced greater reduction in IL-2 levels over the 5-week trial. There was no significant effect of treatment on change in levels of AGA IgG (*p*=0.172) or any components of the standard laboratory tests (CBC, CMP).

#### Side Effects and Adverse Events

There was no significant change in overall side effect burden from baseline to study endpoint on the SAS (*p*=0.78) or BAS (*p*=0.37). Vital sign means were in the normal range and did not differ between groups. There was a significant effect of treatment on change in albumin levels (*df*=29, *F*=4.4, *p*=0.045), such that participants treated with GFD experienced greater increase in albumin levels over the 5-week trial. The occurrence (N, treatment group) of spontaneously reported adverse events were as follows: new onset diabetes mellitus (N=1, GFD), knee pain increase (N=2, GFD), skin lesions (N=1, GFD), acid taste in mouth (N=1, GFD), stomach bloating (N=1, gluten containing), somatic concern (N=1, gluten containing), Covid 19 infection (N=2, GFD).

## Discussion

To our knowledge, this study is the first large-scale confirmatory double-blind randomized controlled trial using a GFD in persons with SRD. It is also the third GFD study from our group (47, 48) showing reduction of negative symptoms in three separate cohorts. In this double-blind randomized controlled trial, we studied the efficacy of GFD in reducing negative symptoms of SRD in persons with high AGA IgG. We found comparatively greater improvement in negative symptoms for persons treated with a GFD over five weeks; specifically, in the ‘apathy/reduced motivation’ domains measured by the CAINS-MAP (*df*=30.1, *F*=2.78, *p*=0.045) and SANS-AA (*df*=29.5, *F*=1.84, *p*=0.15). In contrast, GFD had no effect on negative symptoms in the ‘expressive’ domains (i.e. blunted affect, alogia), depression, or cognitive function. There was a trend for improvement in positive symptoms, but this did not reach significance (p=0.08).

Of note, the ‘apathy/reduced motivation’ domain (i.e., asociality, avolition, anhedonia) is known to have a greater impact on real-world functioning than those negative symptoms included in the ‘expressive’ domain (89-98). We note, however, that, while our finding was significant in reducing anhedonia and avolition in the GFD group compared to the control group, our effect was modest and lower than we have seen previously. Our study was only a 5-week evaluation and likely longer periods will lead to better improvement. Also, this study took place during the COVID-19 pandemic with frequent quarantines and lockdowns, thus impeding our ability to show as robust an effect as we have seen before. Also, of note, our present finding supports the idea of a distinct treatment response in an SRD group with enduring and primary negative symptoms that are not driven by other domains of psychopathology. It also provides further evidence of the connection between this so-called “deficit syndrome” marked by primary and enduring negative symptoms found in about 20% SRD patients (101), which may map onto the same high inflammatory AGA IgG subtype. (102, 103). Nonetheless, these significant results along with positive findings in 2 previous studies supports the use of a GFD in helping treat negative symptomatology in the 33-38% of persons with SRD who have high AGA IgG. This therapeutic benefit is crucial since negative symptoms are a major determinant of social and occupational function (66) and the most significant predictor of poor long-term one-year outcomes (67).

It is important to note that the GFD was well-tolerated in this inpatient study, based on side-effects occurring comparably between groups and only one person discontinuing participation (citing ‘not enough food’ as reason, despite caloric content in the two diets are standardized).

The primary limitation of the study was the COVID-19 pandemic, which involved quarantines, lockdowns, periods of isolation and fearfulness – a nearly insurmountable challenge to the study of social and motivational behaviors. As a form of systematic error, these unpredictable circumstances may have naturally led to behaviors that resembled negative symptomatology, uniformly across treatment groups and time such that our ability to detect greater reductions in negative symptoms and treatment response was hampered.

Similarly, other research groups reported higher negative symptoms, such as anhedonia and avolition, in patients during the pandemic compared to pre-pandemic (104), while other groups found higher anxiety and depression that could contribute to social isolation (105)‥

Our study was also limited by its short five-week design. The short design was based on the rapid changes we observed in our two-week pilot study and the feasibility challenges of an inpatient trial greater than five-weeks. Nonetheless, it may have limited our ability to detect changes in more widespread inflammatory cytokines. AGA IgG antibodies have a half-life greater than five weeks thus, we do not expect this to change but would expect potentially greater improvement after a few months when the antibody levels resume in lower ranges. In addition, we did not control for antipsychotic use, although AGA IgG antibodies are not impacted by antipsychotics the nutritional content of meals do not affect the bioavailability of antipsychotics, and, in general, gluten-containing foods do not affect metabolism of specific antipsychotic medications (106).

In conclusion, our study provides evidence that clinical measures of negative symptomatology and biological markers of inflammation improved with removal of gluten for five weeks in this AGA IgG+ SRD subgroup. This, along with much other accumulating evidence, shows that the deconstruction of schizophrenia may be extremely important for delineating and searching for new treatments (107).

## Data Availability

All data produced in the present study are available upon reasonable request to the authors

## Acknowledgements

The authors would like to thank the participants and their families, without such support this study would not have happened. We would also like to thank the dedicated research staff and nurses at the Spring Grove Hospital Treatment Research Unit who helped to initiate, recruit, organize, rate and operate the clinical trial. Their work, particularly during COVID-19, was commendable and we appreciate all the effort. We particularly acknowledge David Gorelick, MD, PhD, Sharon Pugh PA-C, and Joshua Chiapelli, MD for work on medical clearance and laboratory evaluation and to Daniela Cihakova, PhD for running all AGA and cytokine testing in her laboratory at Johns Hopkins University. We thank Donna Dadkhoo, BS, David Enzana, BS, Hannah Lemke, BA, Bruce J. Patterson, CPT, Megan Powell, MPS, Haley Demyanovich, MPS, Matthew Glassman, MA, Ann Marie Kearns, MS, M. Pat Ball, RN, and Deborah Geisler, MA for their many contributions to the research conduct. Lastly, we are grateful for the support of the analysis team working on this project: Frank Gaston, LCPC, Fang Liu, MS, and Hongji Chen, MS, PhD. Many other investigators should be thanked such as Robert W Buchanan, MD, Stephanie Hare, PhD, Laura Rowland, PhD, Peter Kochunov, PhD, Bhim Adakari, PhD, Gopal Vyas, DO, Heather Adams, PhD, James Waltz, PhD as they played a role in many aspects of this project yet to be published

